# Improved Xerostomia Prediction in Head and Neck Cancer Patients with Dixon Magnetic Resonance Imaging of Glandular Adiposity: Validation of Semi-Quantitative Parotid T1 Signal Intensity Metrics for Biomarker Pre-Qualification

**DOI:** 10.1101/2022.07.11.22277439

**Authors:** Joint Head and Neck Radiotherapy-MRI Development Cooperative, the MD Anderson Head and Neck Cancer Symptom Working Group, Keith L. Sanders, Sam Mulder, Kareem A. Wahid, Brigid A. McDonald, Sara Ahmed, Travis C. Salzillo, Renjie He, Mohamed A. Naser, Cem Dede, Vivian Salama, Ashley Way, Christina Setareh Sharafi, Abdallah S.R. Mohamed, Jillian Rigert, Mark Chambers, Amy C. Moreno, Katherine A. Hutcheson, Stephen Y. Lai, Clifton D. Fuller, Lisanne V. van Dijk

## Abstract

**Purpose:** Parotid whole-gland magnetic resonance (MR) T1 intensity, thresholded at the 90th percentile (T1 P90), has been previously reported to be a candidate MR imaging biomarker (MR-IBM) for improved prediction of xerostomia development after radiotherapy. Although P90 was previously derived from the parotid glands of T1-weighted MRI, in this study, we aim to validate P90 in an external cohort using fat only images reconstructed from a T1 Dixon MRI sequence, as well as determining alternative T1 intensity thresholds for potential qualification as predictive FDA BEST biomarkers of xerostomia development 6 months after radiotherapy (Xero_6m_).

**Methods:** MR-IBMs derived from T1 Dixon intensity-normalized scans from 76 head and neck cancer (HNC) patients were extracted from pre-treatment MR images. Scans were normalized to fat tissue, and imaging characteristics were quantified. A reference model and MR-IBM models were created using multivariable logistic regression to predict Xero_6m_. External validation was performed using the model coefficients described in a previous study. The area under the curve (AUC) of the resulting models were compared. Stepwise forward feature selection was performed to discover additional MR-IBMs for improved predictions of xerostomia.

**Results:** The external validation of a previous model coefficients against our cohort showed decreased performance of the P90 MR-IBM model (AUC of 0.73 (CI 0.61-0.85)). The reference model exhibited improved performance when P90 was incorporated (AUC of 0.78 (CI 0.67-0.89)). Feature selection demonstrated the P10 MR-IBM provided performance improvements (AUC of 0.79 (CI: 0.69-0.90)).

**Conclusion:** Our findings validated P90 as predictive biomarker for radiation-induced xerostomia and showed MR-IBMs derived from Dixon sequences can improve Xero_6m_ prediction when compared to the reference model. Formal biomarker qualification should be considered for T1 sequences/relaxometry via formalized approaches.

## Introduction

In head and neck cancer (HNC) therapy, radiation damage to salivary glands, including the parotid glands, often results in salivary gland hypofunction and resultant xerostomia (dry mouth), associated with long-term, negative impacts on the patient oral health and quality of life [1,2]. Normal tissue complication probability (NTCP) models have been established to estimate the risk of developing radiation-induced xerostomia. Previous studies have constructed NTCP models using the relationship between the radiation dose to the parotid glands and baseline patient-reported xerostomia complaints to predict post-radiotherapy (RT) xerostomia outcomes [3,4]. While these studies have shown potential for predicting xerostomia, there is still unexplained variance in patients who receive similar radiation doses but have different toxicity outcomes. Thus, improving the prediction of radiation-induced xerostomia will assist in the advancement of personalized treatment options through more novel radiation methods such as MR-guided adaptive radiotherapy (ART) [5-7] and proton radiotherapy [8-11].

Biomarkers quantified from medical images i.e., imaging biomarkers (IBMs) can represent the intensity, textural, and geometric characteristics of a region of interest [12]. Utilizing IBMs has demonstrated advantages in improving tumor classification and assessment of patient treatment response [13]. IBMs derived from normal tissue have also been explored for their potential to predict radiotherapy-related toxicities. [14-15]. Specific studies have investigated the role in which the ratio of fat-to-functional parenchymal tissue in the parotid glands can be quantified through IBMs and subsequently used as variables for predicting post-RT xerostomia development [16,17]. While the aforementioned studies utilized CT and PET images, interest in developing magnetic resonance imaging biomarkers (MR-IBMs) has grown due to MRI’s exceptional soft tissue contrast properties.

Previously, van Dijk et al. investigated T1-weighted pre-treatment MR-IBMs of the parotid glands to improve the prediction of late radiation-induced xerostomia [18]. The results of this study identified the 90th intensity percentile (P90) of normalized whole gland T1W intensity to be an effective predictor of xerostomia. However, unlike direct relaxometric T1-mapping, T1-weighted MR acquisitions are not inherently quantitative representations. Our prior hypothesis, that MR-derived fat proportion is a surrogate for xerostomia risk post-RT, suggests that semi-quantitative techniques might provide more accurate measures of parotid composition.

The original method developed by Dixon of “simple proton spectroscopic imaging” leverages constructive interference to derive proportional “in-phase” and “out of phase” images that can then be used to derive “water only” or “fat only” images. The Dixon technique thus allows semi-quantitative measurement of T1-intensity weighted proportional fat content. Dixon estimation of fat content has been widely used in other anatomic sites; our goal in the current study was to determine whether the previously identified T1W P90 held discriminative in the context of a semi-quantitative scan. The purpose of this study was to validate T1W P90 for pre-treatment prediction of patient-rated moderate-to-severe xerostomia development 6 months after treatment (Xero_6m_) when used with mean parotid gland dose and baseline xerostomia complaints, as well as exploring whether other MR-IBMs could provide additional classificatory utility. To undertake this effort, we executed the following specific aims:

1. Evaluate the external validity of T1 P90 MR-IBM by external validation using the model characteristics described by van Dijk et al. on our independent cohort in DIXON MR scans.
2. Determine the model performance of the Dixon P90 MR-IBM refitted to our independent cohort, to assess if the prediction performance of the reference model improved when incorporating the Dixon P90 MR-IBM.
3. Identify alternative or additive MR-IBMs suitable for improving Xero_6m_ prediction for potential biomarker qualification.

## Materials and Methods

### Patient Demographics and Treatment

The study cohort is a secondary analysis of a prospective observational trial cohort. The cohort is comprised of HNC patients treated at MD Anderson Cancer Center (MDACC) between May 2017 and November 2020. Patients were extracted from a prospective imaging/toxicity cohort under secondary use approval from the MD Anderson Cancer Center under IRB RCR03-0800, using data collected under protocol PA16-0302, “Using Magnetic Resonance Imaging (MRI) to Assess Mandibular and Soft Tissue Responses to Radiation Therapy” (NCT03145077). Primary trial eligibility included: patients receiving radiotherapy for head and neck cancer with a minimum dose to the mandible of >= 50Gy. Patients selected for secondary analysis included those who underwent treatment with radiotherapy with or without chemotherapy for curative intent and had no prior radiotherapy treatment, chemotherapy, or HNC surgery. Patients were treated with Intensity-Modulated Radiation Therapy (IMRT) or Volumetric Arc Therapy (VMAT). Patients were prescribed 60-70 Gy of therapeutic dose to tumor volumes over 30-35 fractions delivered over 6-7 weeks. Treatment planning prioritized sparing the parotid glands in the absence of loss of tumor coverage, with a primary dose constraint of <=26 Gy mean dose to the contralateral parotid, parotid V15<30%, and constraint of dose to ipsilateral and contralateral parotid otherwise “as low as reasonably achievable” given target volume location. Patients without pre-treatment or 6 months follow-up data were also excluded. All secondary image registration and data collection was undertaken under HIPPA-compliant protocol (RCR03-0800) with waiver of informed consent approved by The University of Texas MD Anderson Cancer Center Institutional Review Board.

### Endpoints

The primary endpoint was patient-rated moderate-to-severe xerostomia 6 months after radiotherapy (Xero6m). Xero6m was evaluated using the MD Anderson Symptom Inventory (MDASI-HN) questionnaire as part of the Standard Follow-up Program [19]. Symptoms were assessed on a scale from 0-10, with 0 indicating symptom absence and 10 indicating the highest severity of the symptom. In accordance with the MDASI user guide and the purpose of our study, moderate-to-severe xerostomia was defined as MDASI-HN xerostomia score of 5 and higher.

### MRI acquisition and standardization

3D T1-weighted Dixon images were acquired on a 1.5 T clinical MR-sim scanner (Magnetom Aera, Siemens Healthineers, Erlangen, Germany) with the following parameters: FA = 10°, TR/TE = 7.11/2.39,4.77 ms, Pixel Bandwidth = 405 Hz/Pixel, FOV = 256×256 mm^2^, Matrix = 256×256, Slice Thickness = 1 mm, NEX = 2. The in-phase and opposed-phase images were linearly combined to form water only and fat only images.

Fat only Dixon scans were normalized to standardize the MR intensities between patients using cheek fat as a reference structure. First, at least 4 slices of the patient’s subcutaneous fat from the left and right sides of their cheeks at the level of the parotid glands were delineated. To stay consistent with the normalization process previously described by van Dijk et al [18], the Fat-only Dixon images were then multiplied by a set value of 350 and then divided by the average subcutaneous fat intensity value.

### Candidate Predictors

Dose-volume histograms parameters were extracted from the parotid glands, which were segmented on the clinical planning CT. The mean dose delivered to parotid glands (PG Dose) was calculated and used in the subsequent models for estimation of generalized equivalent uniform dose (gEUD).

The parotid glands were manually delineated on pre-treatment Dixon fat images. Delineations were observed by a radiation oncologist who had 8+ years of experience. The “Image biomarker standardization initiative” [20] criteria were used to define the definition and nomenclature of the MR-IBMs extracted from the parotid glands. Using MATLAB 2020a [21], a total of 105 intensity and textural MR-IBMs was extracted. Intensity MR-IBMs described the first-order features of the MR-intensity values, including the mean, median, minimum, maximum and root mean square. Textural MR-IBMs features describe heterogeneity between voxels by involving the spatial distance between gray level MR intensities. Classified as the grey level co-occurrence matrix (GLCM) and the grey level run-length matrix (GLRLM), these second-order features quantify different directional gray level voxel pairs [22] and the number of successive voxels with equivalent gray level [23] respectively.

### Multivariable Analysis and Model Performance

The statistical computing software, R was employed to develop the multivariable logistic regression models and conduct performance analyses [24]. The Regression Modeling Strategies package [25], an extension of the R programming language was used to provide a standardized and well-documented method for model building. The performance metrics evaluated were the area under the ROC (receiver operating characteristic) curve (AUC) and Nagelkerke’s R. The Likelihood-ratio test was used to assess whether an MR-IBM model was significantly different from the reference model.

### Reference Model

A multivariable logistic regression reference model based on the mean PG Dose and baseline xerostomia complaints (Xero_Baseline_) was fitted in the study cohort. Xero_Baseline_ was defined by the MDASI-HN xerostomia score from patients prior to treatment, dichotomized using a scoring threshold of 2, i.e., scores < 2 and scores >=2 corresponding to none and any, respectively.

### External Validation of P90 model

External validation was performed on models fitted to the study cohort to determine if P90 could act as a reproducible and generalizable in xerostomia predictions. The reference (variables: Xero_Baseline_, mean PG dose) and P90 model (variables: Xero_Baseline_, mean PG dose and P90) were externally validated using the corrected coefficients established by van Dijk et al. [18]. Model characteristics used in external validation are described in Table 2.

### Model Update: Refit P90 model

A model update was performed to refit the P90 MR-IBM on the study cohort. The statistical significance of the model coefficients was evaluated with a p-value <0.05. Furthermore, the performance metrics of the refit P90 model were compared to the reference model to evaluate changes in model performance.

### Model Variable Update: Feature Selection

Feature selection was performed to discover additional MR-IBMs which could be effective in xerostomia prediction. To reduce the effects of overfitting and multicollinearity, pre-selection based on Pearson correlation was performed. To stay consistent with similar studies [14,16,18], if the correlation between two candidate MR-IBMs was greater than 0.80, the variable with the highest association with Xero_6m_ was chosen.

Stepwise forward selection was used to create a multivariable logistic regression model based on mean parotid gland dose, Xero_baseline_, and all derived MR-IBMs. The statistical significance of the model coefficients was evaluated with a p-value <0.05. Stepwise forward feature selection was based on the largest significant log-likelihood.

## Results

A total of 76 patients were used for the study cohort. The demographics of the study cohort is described in Table 1. Most of the patients had oropharyngeal carcinomas (78%). A small number of patients reported moderate-to-severe xerostomia at baseline (5%). However, moderate-to-severe xerostomia reports significantly increased 6 months after treatment (36%). In addition, the average (±standard deviation) of mean contralateral PG dose was 16.34 ±13.2 Gy.

**Table 1.** Patient Demographics

| CHARACTERISTICS | N = 76 | % |
| --- | --- | --- |
| SEX |  |  |
| FEMALE | 9 | 11 |
| MALE | 67 | 89 |
| MEDIAN AGE<br>(IQR±) | | |
| AGE | 58 (51.3 - 65) |  |
| TUMOR SITE |  |  |
| LARYNX | 2 | 3 |
| NASOPHARYNX | 6 | 8 |
| ORAL CAVITY | 3 | 4 |
| ORAL TONGUE | 2 | 3 |
| OROPHARYNX | 59 | 78 |
| UNKOWN | 4 | 4 |
| TUMOR CLASSIFICATION |  |  |
| T0 | 6 | 8 |
| T1 | 20 | 26 |
| T2 | 25 | 35 |
| T3 | 10 | 13 |
| T4 | 12 | 15 |
| NODE CLASSIFICATION |  |  |
| N0 | 8 | 10 |
| N1 | 37 | 50 |
| N2A | 4 | 5 |
| N2B | 16 | 21 |
| N2C | 4 | 5 |
| N3 | 3 | 4 |
| NX | 4 | 5 |
| TREATMENT TECHNIQUE |  |  |
| VMAT | 9 | 12 |
| IMPT | 25 | 32 |
| IMRT | 42 | 56 |
| XERO<sub>BASELINE</sub> |  |  |
| YES | 4 | 5 |
| NO | 72 | 95 |

Three evaluations were used to measure the effectiveness of the P90 MR-IBM. The coefficients of all models created during each evaluation are depicted in Table 2. The performance metrics of each evaluation are shown in Table 3.

**Table 2.**
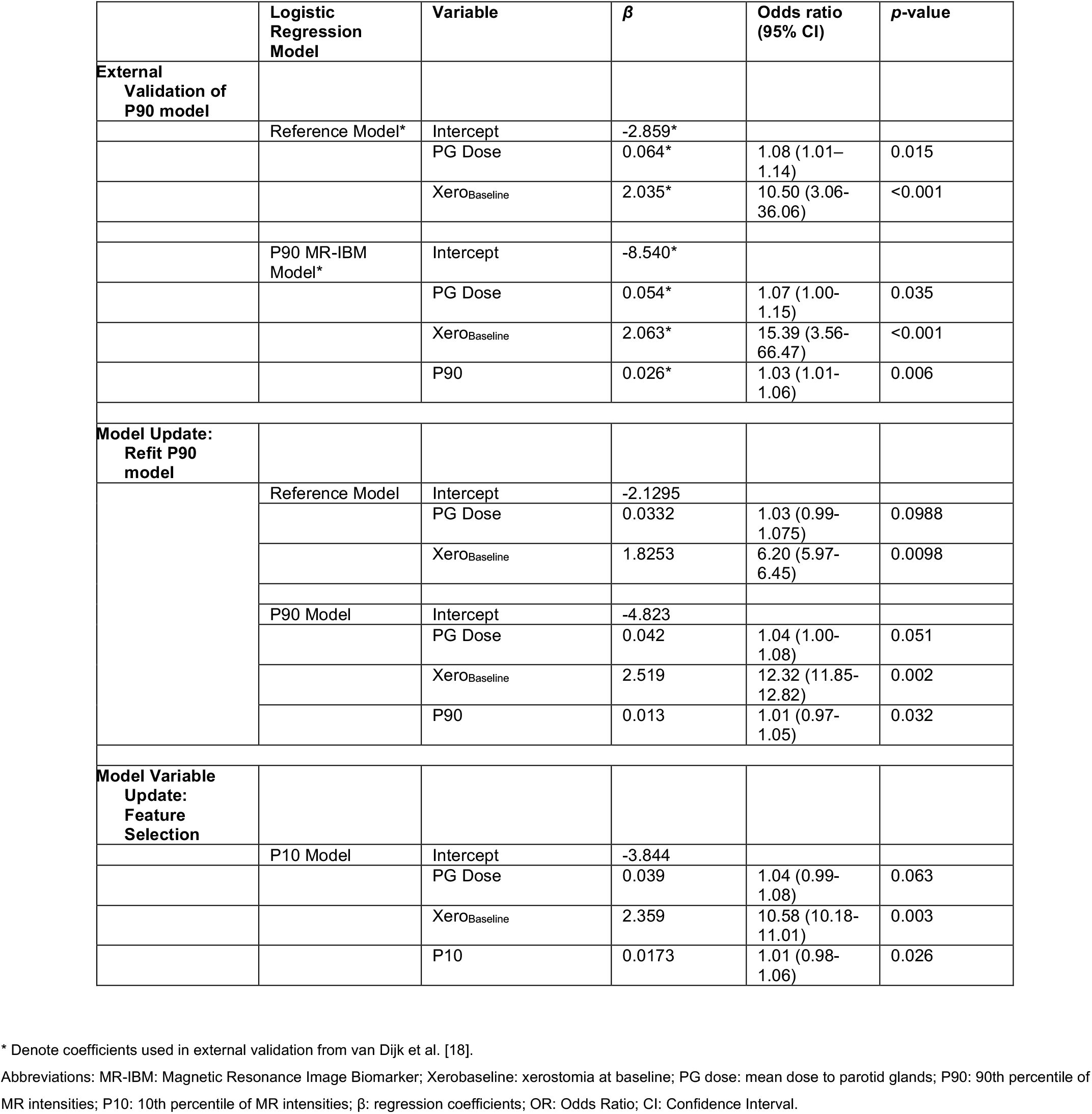
Model characteristics of logistic regression models from external validation, model refit, and variable update evaluations

**Table 3.** Performance metrics of predictive models

|  |  | External Validation* |  | Model Update: Refit P90 model |  | Model Variable Update: Feature Selection |  |
| --- | --- | --- | --- | --- | --- | --- | --- |
|  |  | Reference Model | P90 Model | Reference Model | P90 Model | Reference Model | P10 Model |
| Overall | Nagelkerke's R<sup>2</sup> | 0.17 | 0.19 | 0.18 | 0.26 | 0.18 | 0.26 |
|  | Likelihood Ratio Test | - | - | - | 5.04<br>(p = 0.025) | - | 5.24<br>(p = 0.022) |
| Discrimination | AUC | 0.76<br>(0.65-0.88) | 0.73<br>(0.61-0.85) | 0.76<br>(0.65-0.88) | 0.78<br>(0.67-0.90) | 0.76<br>(0.65-0.88) | 0.79<br>(0.69-0.90) |

\* External validation was performed by using coefficients derived from van Dijk et al. [18].

### Reference Model

The reference model based on mean PG dose and Xero_Baseline_ was fitted to the study cohort data. The performance metrics of the reference model depicted an AUC of 0.76((95%CI: 0.65-0.88); and a R^2^ = 0.18).

### External Validation of P90 model

The coefficients used in external validation (external.val) were described by van Dijk et al. [18] and listed in Table 2. While the reference model showed similar performance in AUC, there was a slight decline in the R^2^ metric (AUCexternal.val = 0.76 (95% CI:0.64-0.87), R^2^external.val = 0.17). However, when the P90 model was externally validated, a reduction in the performance of AUC was observed while R^2^ showed a slight increase (AUCexternal.val = 0.73 (95% CI: 0.61-0.85), R^2^external.val = 0.19).

### Model Update: Refit P90 Model

The reference model was updated by refitting the P90 MR-IBM variable to the study cohort. The model refit showed that incorporating the P90 MR-IBM to the reference model provided considerable improvement to the performance measures, by increasing model’s AUC of 0.76 (95%CI: 0.65-0.88; R^2^ = 0.18) to 0.78 (95%CI 0.67-0.89; R^2^ = 0.26). The model refit was shown to be significantly different from the reference model (Likelihood-Ratio test; p = 0.024).

### Model Variable Update: Feature Selection

The results of the stepwise forward selection indicated that the intensity MR-IBM, the 10^th^ intensity percentile (P10) of the standardized MRI-units to the fat tissue contributed the largest improvement to model performance. The positive coefficient indicated that high P10 is associated with a greater risk of developing Xero_6m_. Fig. 1 showcases patients with high and low P10 values.

**Figure 1.**
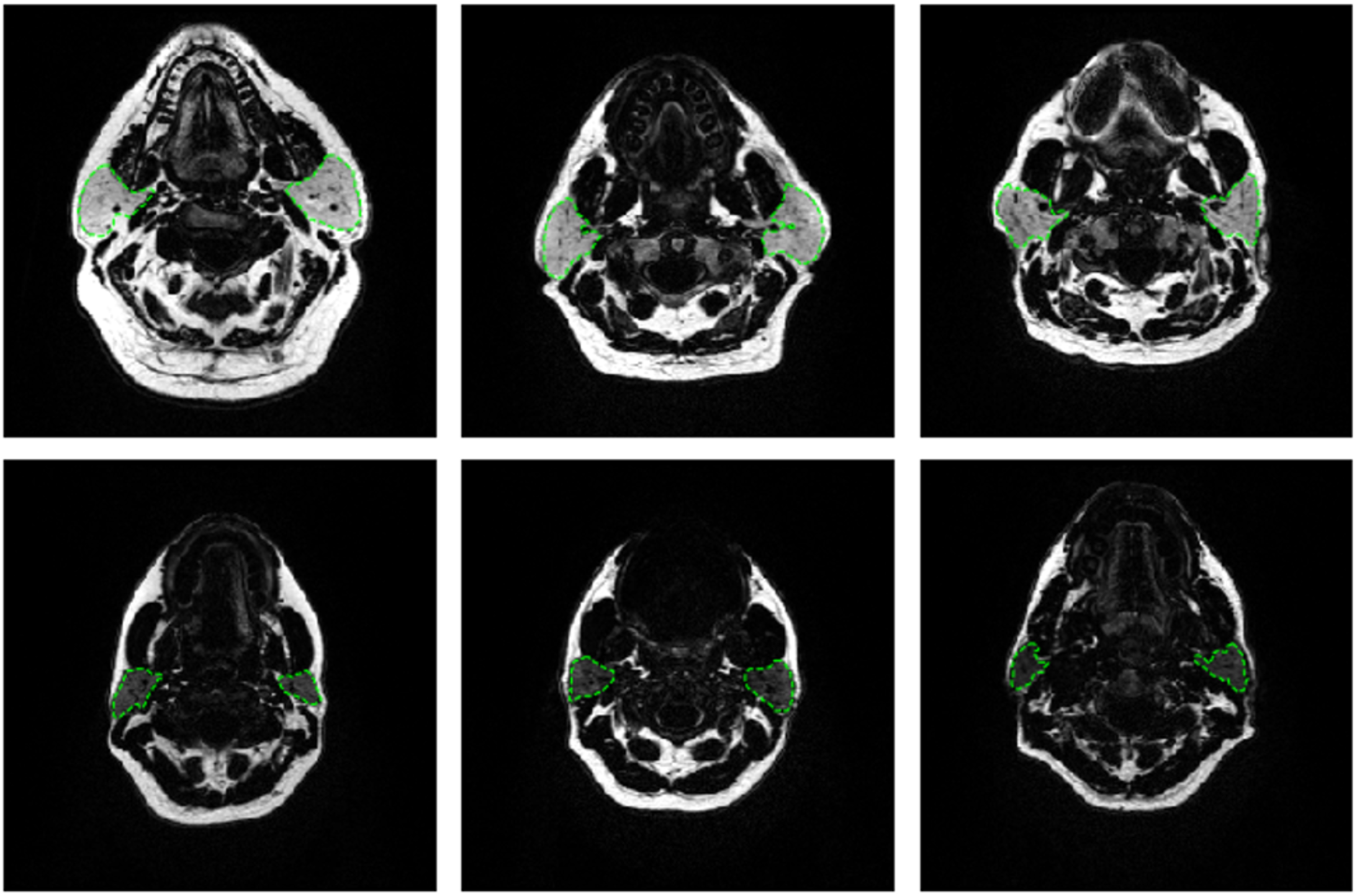
Instances of patients with high (A–C) and low (D–F) P10 values of the parotid glands. Hence, the patients with a higher risk of developing xerostomia 6 months after treatment can be seen on the top row.

The P10 variable was shown to be a significant addition to the reference model the reference model (Likelihood-ratio test; p = 0.021) and resulted in a significant improvement to the performance metric of the reference model, increasing the reference model’s AUC of 0.76 (95%CI: 0.67-0.89; R^2^ = 0.18) to 0.79 (CI: 0.69-0.90), R^2^ = 0.26).

## Discussion

The current study explored the predictive value of 90th percentile of MR-intensities (i.e., P90 MR-IBM) of the parotid glands for late xerostomia in an external data set with fat only images derived from a T1 Dixon sequence. The P90 MR-IBM was previously identified by van Dijk et al. in a T1-weighted TSE sequence [18], indicated the P90 MR-IBM could be used to improve xerostomia prediction 12 months after radiotherapy compared to a reference model, which only utilized mean parotid gland dose and baseline xerostomia complaints. Specifically, higher P90 values were associated with a higher risk of developing xerostomia. Although MR-IBMs have shown promise in improving xerostomia predictions, the wide variety of MRI sequence types introduce an unmet opportunity to validate previously described MR-IBMs in different sequences.

Similar studies have investigated using the fat-to-functional parenchymal parotid tissue ratio, quantified through IBMs as an indicator for radiation-induced xerostomia predictions [16,17]. These studies utilized IBMs derived from CT and ^18^FDG PET imaging modalities, resulting in findings that suggested that IBMs characterizing increased heterogeneous CT intensity and low metabolic ^18^FDG-PET of the parotid glands were associated with a higher risk of xerostomia development 12 months after radiotherapy. The advantage MR provides over CT and PET imaging is that the acquired image offers high soft-tissue contrast without utilizing ionizing radiation. Moreover, when identifying IBMs for improving radiation-induced xerostomia predictions, MR-IBMs have been shown to outperform CT-IBMs. A study performed by Sheikh et al. developed multiple logistic regression models predicting xerostomia 3 months after radiotherapy. The results of the study revealed that incorporating the MR-IBMs of the salivary glands along with dosimetric parameters, provided superior improvement in model performance when compared to models which utilized CT-IBMs [11].

The current study is based on the Dixon technique, which is unique to other fat suppression methods as it can reconstruct uniform fat and water images in post-processing [26]. Taking advantage of their differing chemical shifts, the in-phase and opposed-phase Dixon sequences can be linearly combined to produce images that suppressed water signal, thus assessing fat content semi-quantitatively. While the Dixon sequences have been used to evaluate chronic sacroiliitis and muscular dystrophy [28,29], to our knowledge, this sequence is less explored in radiation-induced xerostomia. A study conducted by Rosen et al. illustrated the use of fat fraction changes in the parotid gland measured by Dixon imaging during the beginning stages of radiotherapy as a viable variable for improving acute xerostomia prediction [30]. Since the Dixon fat only images are superior to T1-weighted images in fat quantification [27], we hypothesize that the P90 MR-IBM derived from pre-treatment T1 Dixon fat scans can be used to investigate the relationship between fat-to-functional parotid tissue and radiation-induced xerostomia by providing improvements to the performance metrics of post-RT xerostomia prediction models. The results of our study demonstrated improvements in model performance when the P90 MR-IBM was incorporated into the model, thus confirming our hypothesis. Furthermore, our study highlighted the capabilities of intensity MR-IBMs as a predictor variable by suggesting the P10 MR-IBM through feature selection.

Three evaluations were performed to investigate whether the P90 MR-IBM was also predictive in an independent cohort with a similar, yet distinct, T1 Dixon MR sequence. Our first evaluation showed that the recently published MR-IBM NTCP model [18] that was developed on T1-TSE MR scans, could not be directly validated in our cohort with the P90 MR-IBM extracted from the T1 Dixon fat images. An important distinction in the purpose of this evaluation was to validate the P90 MR-IBM described by van Dijk et al. and not directly validate the prediction model. The external validation results of the current study’s reference model against the reference model coefficients described by van Dijk et al. [18] revealed similar performance metrics. This suggests that the reference models based on mean parotid gland dose and baseline xerostomia complaints are generalizable across the different study populations. The decrease in the performance metrics of the reference model (AUCexternal.val = 0.76) compared to the P90 model (AUCexternal.val = 0.73) suggests that the P90 MR-IBM may not perform as well across study populations. The decrease in performance could be explained by how P90 is assessed in the different MR sequences. While the differences may have had a detrimental effect on the validation evaluation, the goal of the study was achieved by the refit and feature selection evaluations respectively.

The results of our second evaluation, the model update showed that P90 alone was not significantly associated with Xero_6m_. However, P90 was significantly associated with Xero_6m_ when used in conjunction with the mean parotid dose and baseline complaints. The addition of the P90 MR-IBM improved the performance measure of the reference model (AUC of 0.76 increased to 0.78). The findings suggest that P90 can be used as a pre-treatment MR-IBM to improve the prediction of radiation-induced xerostomia. Furthermore, our results complement previous studies by also demonstrating the fat-to-functional parenchymal parotid tissue ratio as an indicator for radiation-induced xerostomia predictions [16-18].

Stepwise forward selection showed the intensity MR-IBM P10 as the variable which provided the most significant improvement to model performance. P10 indicates the 10th percentile of the MR-intensities of the parotid glands. The model coefficients suggest that higher P10 values were associated with a higher risk of developing xerostomia. Univariable analysis showed that P10 alone did not show a significant association with Xero_6m_. Nevertheless, P10 was significantly associated with Xero_6m_ when used in combination with mean parotid dose and baseline complaints. Incorporating P10 improved the performance measure of the reference model (AUC increased from 0.76 to 0.79). Since P10 is used as a surrogate variable to assess the predisposed fat content of the parotid gland, these findings indicate that fat content alone does not improve radiation-induced xerostomia predictions, thus the involvement of clinical and dosimetric information is still necessary.

The PG Dose variable, indicating the mean dose to the parotid glands was not significantly associated with Xero_6m_. Furthermore, PG Dose was not a statistically significant variable included in the reference and MR-IBM models (both P90 and P10 models). However, these finding reinforces our need to explore radiation-induced xerostomia toxicity prediction by demonstrating radiation dose delivered to the parotid glands may not explain the variation among patients who develop radiation-induced xerostomia.

Although the performance metrics show higher AUC improvement when incorporating P10 compared to the P90 (AUC 0.79 and AUC 0.78 respectively) until a validation study is performed our findings cannot indicate P10 as a superior variable for improving radiation-induced xerostomia predictions. While the P90 and P10 MR-IBMs both measure the MR-intensity percentiles, they are fundamentally different as they represent an intensity value higher than 90% and higher than 10% of the parotid glands respectively. The P90 MR-IBM suggest that patients are associated with a higher risk of developing radiation-induced xerostomia if 10% of their parotid gland volume contains high-intensity values. Meanwhile, the P10 MR-IBM suggest that patients are associated with a higher risk of developing radiation-induced xerostomia if 90% of their parotid glands contain high-intensity values. Due to the normalization process during the MRI standardization, both P90 and P10 MR-IBMs still suggest that higher intensity values are associated with a higher fat concentration in the parotid glands and thus a higher risk of developing xerostomia 6 months after treatment. Furthermore, our findings complement the findings of van Dijk’s study [18], by demonstrating that intensity-based MR-IBMs may be used to improve radiation-induced xerostomia predictions.

There are several differences between the study conducted by van Dijk et al. [18] and our current study which should be discussed. The current study is composed of patients treated in the United States, which has different treatment planning practices than patients used in van Dijk’s study which consisted of European treated patients. Moreover, the grading scales for patient-rated xerostomia scores were different as the current study used a 0-10 scale MDASI-HN questionnaire and van Dijk’s study used a 4-point Likert scale of the EORTC QLQ-H&N35 questionnaire. However, the differences in grading scales seemed to have little impact likely due to the thresholds used to dichotomize groups i.e. baseline xerostomia complaints and postRT xerostomia complaints. An additional difference between the two studies is that the endpoint of the current study evaluated xerostomia development 6 months after treatment and van Dijk et al. evaluated xerostomia development 12 months after treatment. The external validation in the reference model implies that these differences had no impact on the findings of the evaluation. Differences that impacted the results of the external validation were observed in the decreased performance metric validating the P90 MR-IBM model.

A crucial goal of this study was to expand our understanding of how integrating MR-IBMs could improve our predictions of radiation-induced toxicity in HNC patients. While the findings of this study provided supplementary steps, future studies could be incorporated to further achieve this goal. For instance, this study focused on deriving MR-IBMs from the parotid glands however, other salivary secretion glands could provide additional information useful to xerostomia prediction. Moreover, investigating MR-IBMs from additional timepoints i.e. mid-RT and post-RT, and involving subsequent MRI sequences such as DWI and DCE could support the usefulness of MR-IBMs in prediction models.

## Conclusion

In conclusion, this study validated the novel P90 intensity MR-IBM by deriving it from the T1 Dixon fat only MR images and evaluating its inclusion in model performance. Additionally, the results of this study support the hypothesis that a high concentration of fat in the parotid glands, quantified by intensity MR-IBMs, is related to a higher risk of Xero_6m_. The Xero_6m_ prediction model based on mean parotid dose and baseline xerostomia complaints showed performance improvements when incorporating MR-IBM P90, thus complementing the findings of previous studies. Furthermore, our findings showed the P10 intensity MR-IBM to provide further improvement to model performance. While this study provides additional merit to the P90 intensity MR-IBM and the fat-to-functional parotid tissue hypothesis, we hope that our study encourages more exploration in incorporating MR-IBMs into clinical prediction models.

## Supporting information

Supplement: TRIPOD reporting checklist

## Data Availability

Associated data are available as an online repository at figshare.com, pending completion of peer review.

https://doi.org/10.6084/m9.figshare.20278068

---

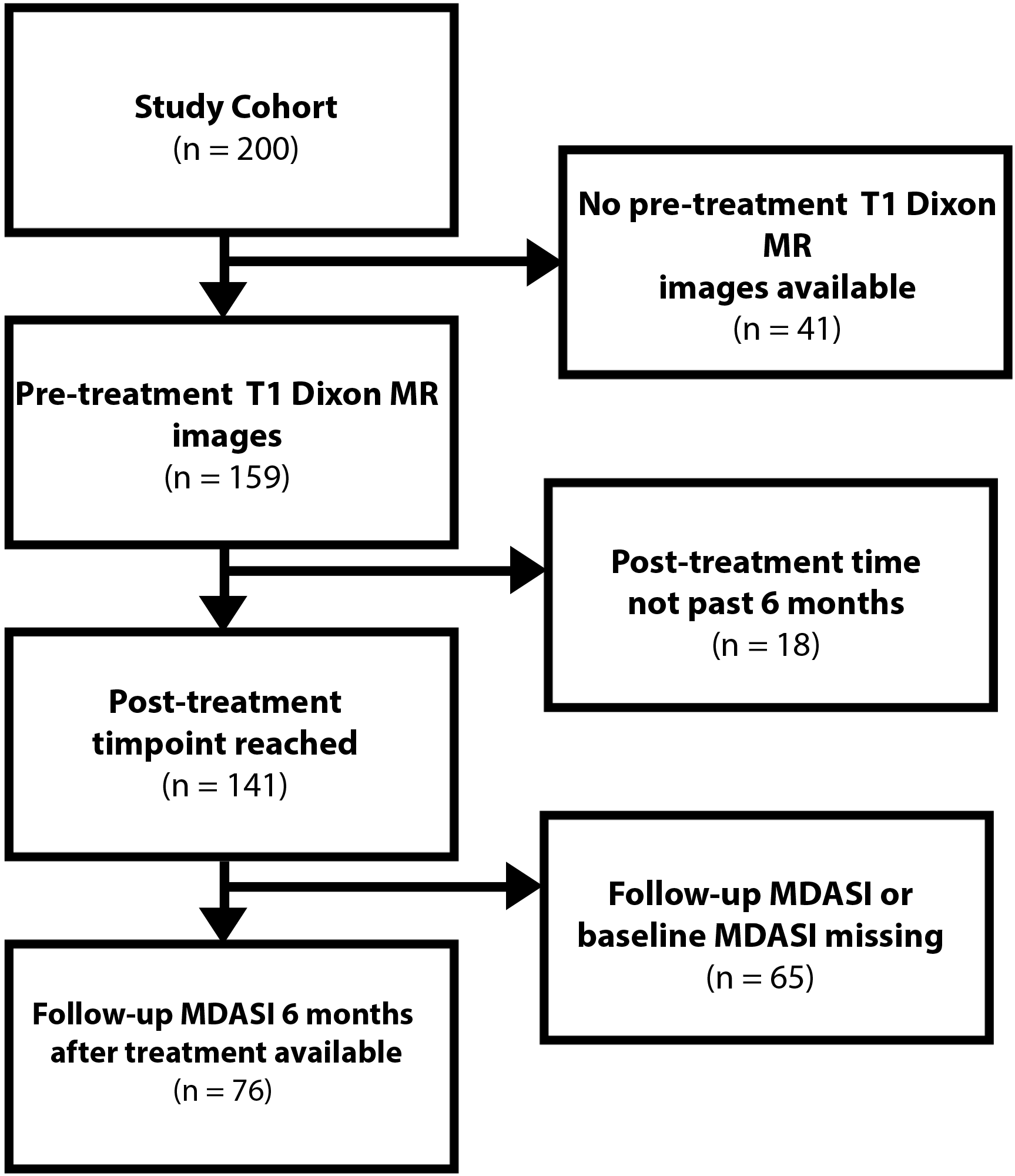

## Notes

<u>Funding/acknowledgements:</u> KLS was supported by the National Institutes of Health (NIH) National Institute for Craniofacial and Dental Research (NIDCR) Research Supplement to Promote Diversity in Health-Related Research (DE028290-01S1).

SM is supported by direct funds from the NIDCR (R01 DE028290).

KAW received/receives support from the Dr. John J. Kopchick Fellowship through The University of Texas MD Anderson UTHealth Graduate School of Biomedical Sciences, the American Legion Auxiliary Fellowship in Cancer Research, The University of Texas Health Science Center at Houston (UTHealth) Center for Clinical and Translational Sciences (CCTS) TL1 Program (TL1TR003169), and an NIH NIDCR Graduate Fellowship (F31DE031502).

BAM received/receives support via the Dr. John J. Kopchick Fellowship, and an NIH NIDCR Graduate Fellowship (F31DE029093).

TCS received/receives funding support from the UTHealth CCTS TL1 Program (TL1TR003169).

CD received/receives funding support from the NIH/National Science Foundation (NSF) National Cancer Institute (NCI) Smart and Connected Health Program (R01CA257814-01).

ASRM received/receives funding and salary support from related to this project from: NIDCR Academic Industrial Partnership Grant (R01DE028290); NIDCR Establishing Outcome Measures for Clinical Studies of Oral and Craniofacial Diseases and Conditions award (R01DE025248); NIH/NSF NCI Smart Connected Health Program (R01CA257814).

AW received direct support from the NCI Small Business Innovation Research Grant Program through a Research Supplement to Promote Diversity (R43CA254559-S1).

JR receives direct support from a NIDCR Research Supplement to Promote Diversity in Health-Related Research (DE028290-S2).

ACM received/receives support from the NCI Paul Calabresi Clinical Trial Program (K12CA088084), a NIDCR NIH Exploratory/Developmental Research Grant Program (R21DE031082), and a NIDCR Mentored Career Development Award to Promote Diversity in the Dental, Oral and Craniofacial Workforce (K01 DE030524).

KAH received/receives support from the Patient-Centered Outcomes Research Institute (PCORI) (PCS-1609-36195) and NCI Clinical and Translational Exploratory/Developmental Studies Grant (R21CA226200).

SYL received/receives support from an NCI Exploratory/Developmental Bioengineering Research Grant (R21CA259839), NIDCR Establishing Outcome Measures for Clinical Studies of Oral and Craniofacial Diseases and Conditions Grant (R01DE025248), and the Ruth L. Kirschstein National Research Service Award (NRSA) Institutional Research Training Grant Program (T32CA261856).

CDF received/receives funding and salary support during the period of study execution from: the NIH National Institute of Biomedical Imaging and Bioengineering (NIBIB) Research Education Programs for Residents and Clinical Fellows Grant (R25EB025787); NIDCR Academic Industrial Partnership Grant (R01DE028290); NCI Early Phase Clinical Trials in Imaging and Image-Guided Interventions Program (1R01CA218148); NCI Institutional Research Training Grant Program (T32CA261856); NIDCR NIH Exploratory/Developmental Research Grant Program (R21DE031082); the NCI SBIR Grant Program (R43CA254559); an NIH/NCI Cancer Center Support Grant (CCSG) Pilot Research Program Award from the UT MD Anderson CCSG Radiation Oncology and Cancer Imaging Program Seed Grant (P30CA016672); a Joint NSF/NIH Initiative on Quantitative Approaches to Biomedical Big Data (QuBBD) Grant (R01CA225190); and an NSF Division of Civil, Mechanical, and Manufacturing Innovation (CMMI) grant (NSF 1933369). CDF has received direct industry grant support, honoraria, and travel funding from Elekta AB unrelated to this project.

SvD was supported by a grant from the Dutch Research Council/Nederlandse Organisatie voor Wetenschappelijk Onderzoek (NWO 452182317).

Direct infrastructure support is provided to by the multidisciplinary the Radiation Oncology/Cancer Imaging Program (P30CA016672-44) of the MD Anderson Cancer Center Support Grant (P30CA016672), the MD Anderson Cancer Center Stiefel Oropharyngeal Research Fund, and the MD Anderson Program in Image-guided Cancer Therapy.

### Competing Interest Statement

CDF has received direct industry grant support, honoraria, and travel funding from Elekta AB unrelated to this project.

### Clinical Protocols

https://clinicaltrials.gov/ct2/show/NCT03145077

### Funding Statement

Funding/acknowledgements:
KLS was supported by the National Institutes of Health (NIH) National Institute for Craniofacial and Dental Research (NIDCR) Research Supplement to Promote Diversity in Health-Related Research (DE028290-01S1).
SM is supported by direct funds from the NIDCR (R01 DE028290).
KAW received/receives support from the Dr. John J. Kopchick Fellowship through The University of Texas MD Anderson UTHealth Graduate School of Biomedical Sciences, the American Legion Auxiliary Fellowship in Cancer Research, The University of Texas Health Science Center at Houston (UTHealth) Center for Clinical and Translational Sciences (CCTS) TL1 Program (TL1TR003169), and an NIH NIDCR Graduate Fellowship (F31DE031502).
BAM received/receives support via the Dr. John J. Kopchick Fellowship, and an NIH NIDCR Graduate Fellowship (F31DE029093).
TCS received/receives funding support from the UTHealth CCTS TL1 Program (TL1TR003169).
CD received/receives funding support from the NIH/National Science Foundation (NSF) National Cancer Institute (NCI) Smart and Connected Health Program (R01CA257814-01).
ASRM received/receives funding and salary support from related to this project from: NIDCR Academic Industrial Partnership Grant (R01DE028290); NIDCR Establishing Outcome Measures for Clinical Studies of Oral and Craniofacial Diseases and Conditions award (R01DE025248); NIH/NSF NCI Smart Connected Health Program (R01CA257814).
AW received direct support from the NCI Small Business Innovation Research Grant Program through a Research Supplement to Promote Diversity (R43CA254559-S1).
JR receives direct support from a NIDCR Research Supplement to Promote Diversity in Health-Related Research (DE028290-S2).
ACM received/receives support from the NCI Paul Calabresi Clinical Trial Program (K12CA088084), a NIDCR NIH Exploratory/Developmental Research Grant Program (R21DE031082), and a NIDCR Mentored Career Development Award to Promote Diversity in the Dental, Oral and Craniofacial Workforce (K01 DE030524).
KAH received/receives support from the Patient-Centered Outcomes Research Institute (PCORI) (PCS-1609-36195) and NCI Clinical and Translational Exploratory/Developmental Studies Grant (R21CA226200).
SYL received/receives support from an NCI Exploratory/Developmental Bioengineering Research Grant (R21CA259839), NIDCR Establishing Outcome Measures for Clinical Studies of Oral and Craniofacial Diseases and Conditions Grant (R01DE025248), and the Ruth L. Kirschstein National Research Service Award (NRSA) Institutional Research Training Grant Program (T32CA261856).
CDF received/receives funding and salary support during the period of study execution from: the NIH National Institute of Biomedical Imaging and Bioengineering (NIBIB) Research Education Programs for Residents and Clinical Fellows Grant (R25EB025787); NIDCR Academic Industrial Partnership Grant (R01DE028290); NCI Early Phase Clinical Trials in Imaging and Image-Guided Interventions Program (1R01CA218148); NCI Institutional Research Training Grant Program (T32CA261856); NIDCR NIH Exploratory/Developmental Research Grant Program (R21DE031082); the NCI SBIR Grant Program (R43CA254559); an NIH/NCI Cancer Center Support Grant (CCSG) Pilot Research Program Award from the UT MD Anderson CCSG Radiation Oncology and Cancer Imaging Program Seed Grant (P30CA016672); a Joint NSF/NIH Initiative on Quantitative Approaches to Biomedical Big Data (QuBBD) Grant (R01CA225190); and an NSF Division of Civil, Mechanical, and Manufacturing Innovation (CMMI) grant (NSF 1933369). CDF has received direct industry grant support, honoraria, and travel funding from Elekta AB unrelated to this project.
LvD was supported by a grant from the Dutch Research Council/Nederlandse Organisatie voor Wetenschappelijk Onderzoek (NWO 452182317) and with a Young Investigator Grant from KWF Dutch Cancer Society (KWF 13529. Direct infrastructure support is provided to by the multidisciplinary the Radiation Oncology/Cancer Imaging Program (P30CA016672-44) of the MD Anderson Cancer Center Support Grant (P30CA016672), the MD Anderson Cancer Center Stiefel Oropharyngeal Research Fund, and the MD Anderson Program in Image-guided Cancer Therapy.

### Author Declarations

The Institutional Review Board of MD Anderson Cancer Center gave ethical approval for this secondary data analysis with waiver of informed consent (RCR03-0800); patients had previously supplied consent to the primary protocol under which data was collected, PA16-0302(NCT03145077).

